# Impact of gastrointestinal comorbidities in patients with right & left atrial isomerism

**DOI:** 10.1101/2020.05.24.20112078

**Authors:** Anastasia Fotaki, Victoria L Doughty, Winston Banya, Stefano Giuliani, Sarah Bradley, Julene S Carvalho

## Abstract

**Objective:** Heterotaxy syndrome (HS), being right atrial isomerism (RAI) or left atrial isomerism (LAI) often presents with congenital heart disease (CHD). Intestinal abnormalities including malrotation are common. We aim to assess the impact of gut abnormalities on outcome in HS.

**Methods:** We reviewed cardiology records of HS patients regarding presence of CHD, time for cardiac intervention, presence of gastrointestinal (GI) abnormalities and type/time of surgery. A questionnaire about GI status was sent to patients <18 years old. Kaplan-Meier curves were derived for survival data.

**Results:** Data were available for 195 patients (49 RAI, 146 LAI) of 247 identified. Questionnaires were sent to 77 families, 47 replied. CHD was present in all RAI and 63.7% of LAI cases. Thirty-eight patients had abdominal surgery (19.5%), similar rate in RAI and LAI (20.4% vs 19.1%, p=0.92). Ladd procedure was performed in 17 (44.7%), non-Ladd in 12 (31.5%) and both procedures in nine patients (23.7%). Ten-year freedom from Ladd procedure for all was 86% (RAI=87%; LAI=85%, p=0.82). Freedom from any GI surgery at one year was 86% (RAI=86%; LAI=86%, p=0.98) and at ten years was 80% (RAI=77%; LAI=81%, p=0.65). Ten-year freedom from cardiac surgery was 34% (RAI=7%; LAI=42%, p<0.0001).

**Conclusions:** In our cohort, one in five patients required abdominal surgery, mostly in their first year, similar in RAI and LAI. Between one and ten years of follow up the impact of GI abnormalities on outcome was minimal. Medium term survival was related to CHD.

**ARTICLE SUMMARY:** *STRENGTHS AND LIMITATIONS OF THIS STUDY:*

- This study is the largest cohort study investigating the impact of gastrointestinal abnormalities in cardiology patients with heterotaxy syndrome.
- It is the first clinical study to show that HS patients suffer from a wider spectrum of abdominal abnormalities, other than typical malrotation, varying in severity from asymptomatic malrotation to complete non-rotation, namely atresia at multiple intestinal levels. This is relevant in prenatal family counseling but also raises questions regarding the indication of elective Ladd procedure in all heterotaxy syndrome patients, as a different procedure might be indicated.
- Patient morbidity was investigated from both the cardiology and gastrointestinal point 12 of view.
- Its main limitations in design is that is not a prospective study. Cardiology records have been reviewed with regards to GI symptomatology, screening investigations and procedures.
- It involved both reviewing the medical records and directly contacting the family for the subcohort that was below 18 years old and lived in the UK, in an attempt to minimise any data errors. Our records were consistent with the information provided by parents/guardians in all cases but one.

## INTRODUCTION

Heterotaxy syndrome (HS) refers to the abnormal arrangement of organs along the left-right axis that is neither situs solitus nor inversus.^1, 2^ Birth incidence is reported as 1:10,000.^3, 4^ HS can be further divided in two diagnostic categories: right atrial isomerism (RAI) and left atrial isomerism (LAI). In LAI there is dominance of left-sided structures, typically associated with multiple spleens (polysplenia), bilateral bilobed lungs, bilateral hyparterial bronchi and two morphological left atria. The inferior vena cava is interrupted with azygos continuation to the superior vena cava (SVC). In right atrial isomerism (RAI), there is dominance of right-sided structures with bilateral trilobed lungs, bilateral eparterial bronchi and two morphological right atria. Asplenia is common and often there are bilateral SVCs.^5^ The cardiac development in heterotaxy is often abnormal and leads to a wide spectrum of cardiac defects. Congenital heart disease (CHD) is common in both LAI and RAI and often more complex in RAI.

HS is often associated with abnormal rotation and fixation of the intestine, termed malrotation. Gut malrotation is thought to occur in 40-90% of patients with HS.^6-11^ The Ladd procedure is the surgical treatment for malrotation with midgut volvulus. Briefly, it consists of de-rotation of the twisted small bowel.^12^ However, no consensus exists regarding whether HS patients should be electively screened for malrotation. It is unclear whether elective, prophylactic Ladd procedure is indicated for asymptomatic patients, especially as their complex cardiac anatomy and cardiorespiratory physiology, predisposes them to adverse effects during the perioperative period. However, improved medical care for children with CHD and HS has increased clinicians’ interest in intestinal comorbidities associated with HS. Additionally, with advances in prenatal screening,^13^ more children with HS and less severe cardiac abnormalities or with a functionally normal heart are being identified before birth. In the latter groups without or with less severe cardiac defects, it is particularly important to know how much the presence of gut abnormalities leading to symptoms and/or surgery can impact on morbidity and mortality in children with HS.

The objective of this research project is to ascertain the impact of gut abnormalities in children with HS to help guide management in the current era of better survival from cardiac surgery and improved prenatal screening.

## METHODS

This is a retrospective study of patients with known RAI or LAI, identified from paediatric and fetal cardiac databases in our tertiary institutions, Royal Brompton Hospital and St George’s University Hospital. All available medical records were reviewed retrospectively regarding the presence and type of abdominal symptoms, screening examinations undertaken, surgical interventions and documentation of any related major complications. Information regarding presence of CHD, need and time for cardiac surgery was also recorded. Outcome (alive or dead) at the time of data review was noted and all live patients under the age of 18 years living in the UK were identified. A letter was posted to the parents of the patients in this group inviting them to complete a questionnaire (Supplementary material 1 and 2) regarding their child’s abdominal symptoms, date and time of any screening test for abdominal problems, date and type of any abdominal surgery and known complications following surgery. Two different questionnaires were designed, which reflected if the child was known to have undergone previous abdominal surgery (Supplementary material 1) or not (Supplementary material 2), based on their medical records.

The study was approved by the National Research Ethics Committee (Reference: 15-SC-0504). The need for informed consent for retrospective review of data was waived, but consent was obtained from those contacted by post.

### Patient and public involvement statement

During prenatal counseling of parents with fetuses with HS, questions about extracardiac comorbidities arose frequently. Especially for the fetuses with LAI and minor heart defects, it was hard to advise on the extracardiac manifestations. Review of the recent literature had a significant gap in the analysis of paediatric cardiology records with regards to GI disease, and thus we undertook this study.

Patients were not involved in the recruitment of the study. However for a subgroup of patients that were alive and below 18, a questionnaire with the study research questions, was posted to their parents. A majority of those answered the questionnaire and reported interest to be informed on the results of the study. This is facilitated by post again.

#### Statistical analysis

Categorical data are presented as number (n, %) and comparisons between groups are made using the chi-squared or Fisher’s exact test. Numeric data are presented as median (inter-quartile range, IQR).

Kaplan-Meier survival curves were plotted to assess freedom from cardiac intervention, any GI surgery and Ladd surgery for the whole cohort and for RAI versus LAI patients. The Logrank test was used to assess difference in freedom from intervention between these two groups. All tests were two-sided and significance was set at p <0.05. The analysis was done using STATA version 14.1.

## RESULTS

A total of 247 patients with the diagnosis of RAI or LAI were identified. Of these, 195 patients were included in this study and constitute our cohort, 52 were excluded from the study due to insufficient information in medical notes (n = 45) or the diagnosis of isomerism was unclear (n = 7). The oldest patient was born in 1947 and the youngest in 2015.

Letters and appropriate questionnaires were posted to patients that were alive and under 18 years old living in the UK (n= 77). Families’ replies (n=45, 58%)were in agreement with the information obtained from the medical notes, in all but one. In this case, the child had had surgery for duodenal atresia and malrotation, which was not reported in the medical notes

Of the 195 cases, 146 patients had LAI (74.9%) and 49 had RAI (25.1%). All cases diagnosed with RAI had CHD, compared to 63.7% of cases with LAI (Table 1). Kaplan-Meier curves indicating time to first cardiac intervention in the whole cohort and comparing those with RAI and LAI are shown in Figures 1a and 1b.

**Figure 1a:**
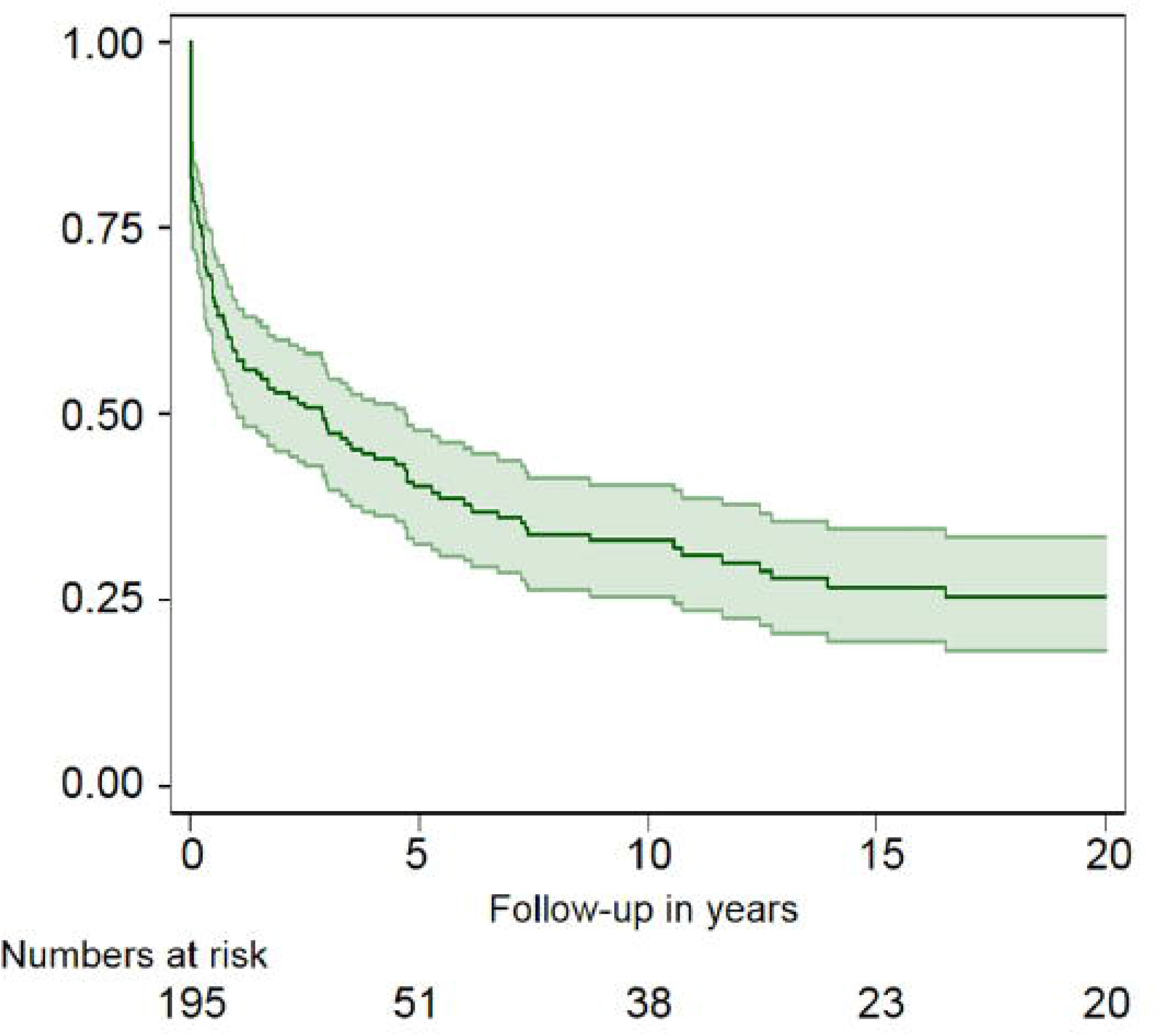
Kaplan-Meier curve, showing freedom from cardiac intervention for the whole cohort.

**Figure 1b:**
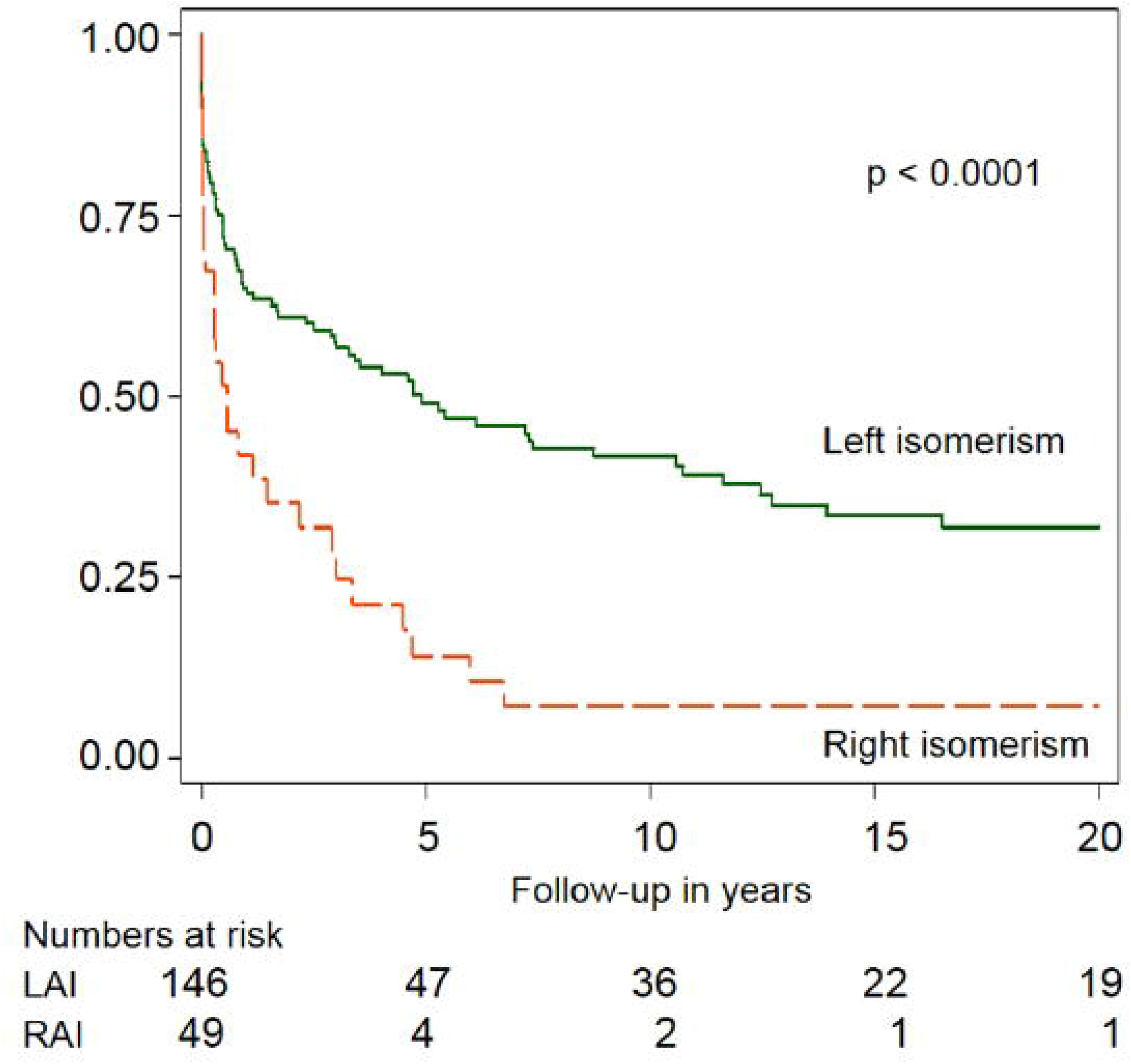
Kaplan-Meier curve, comparing freedom from cardiac intervention for RAI versus LAI.

**Table 1:**
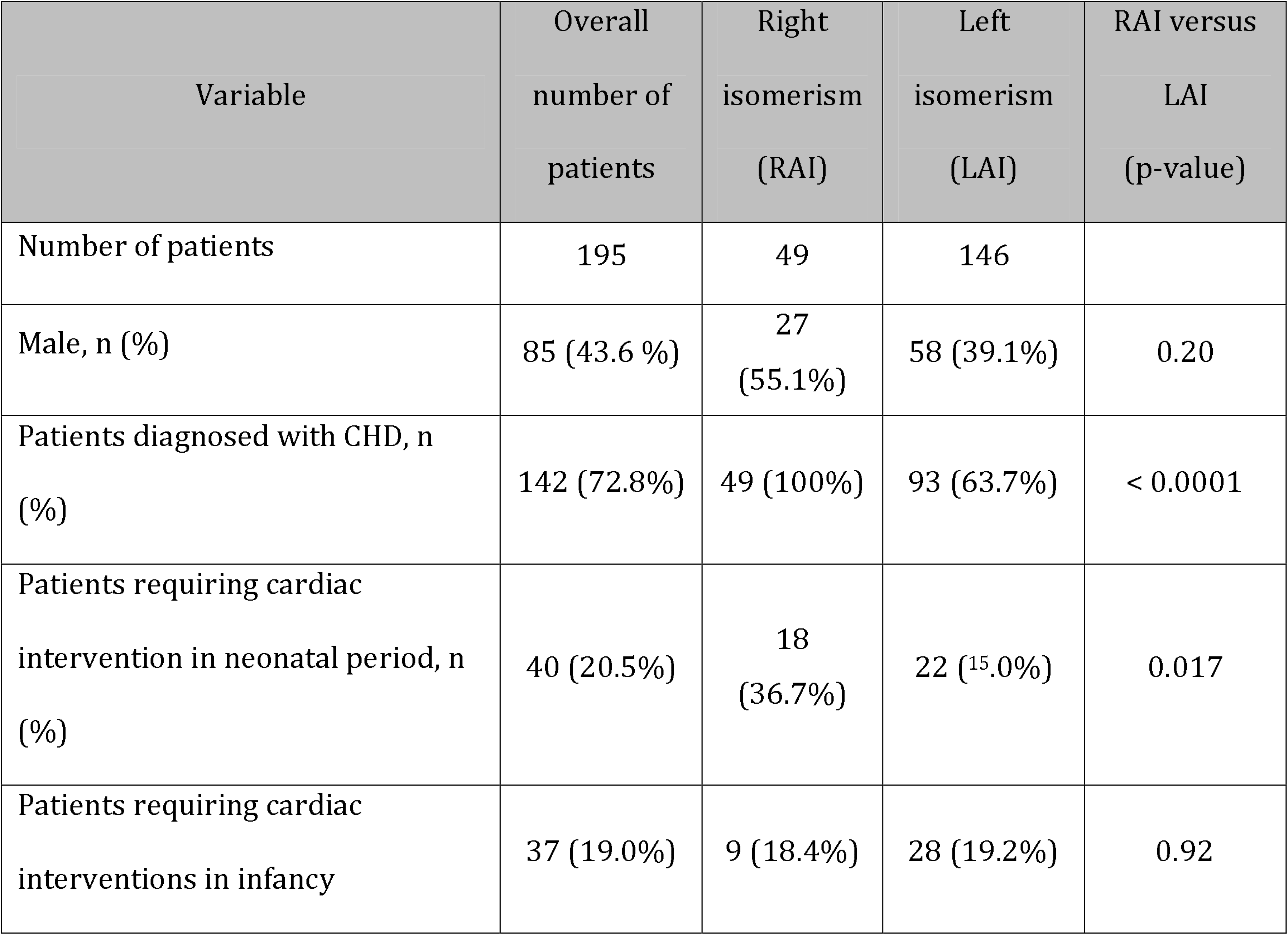
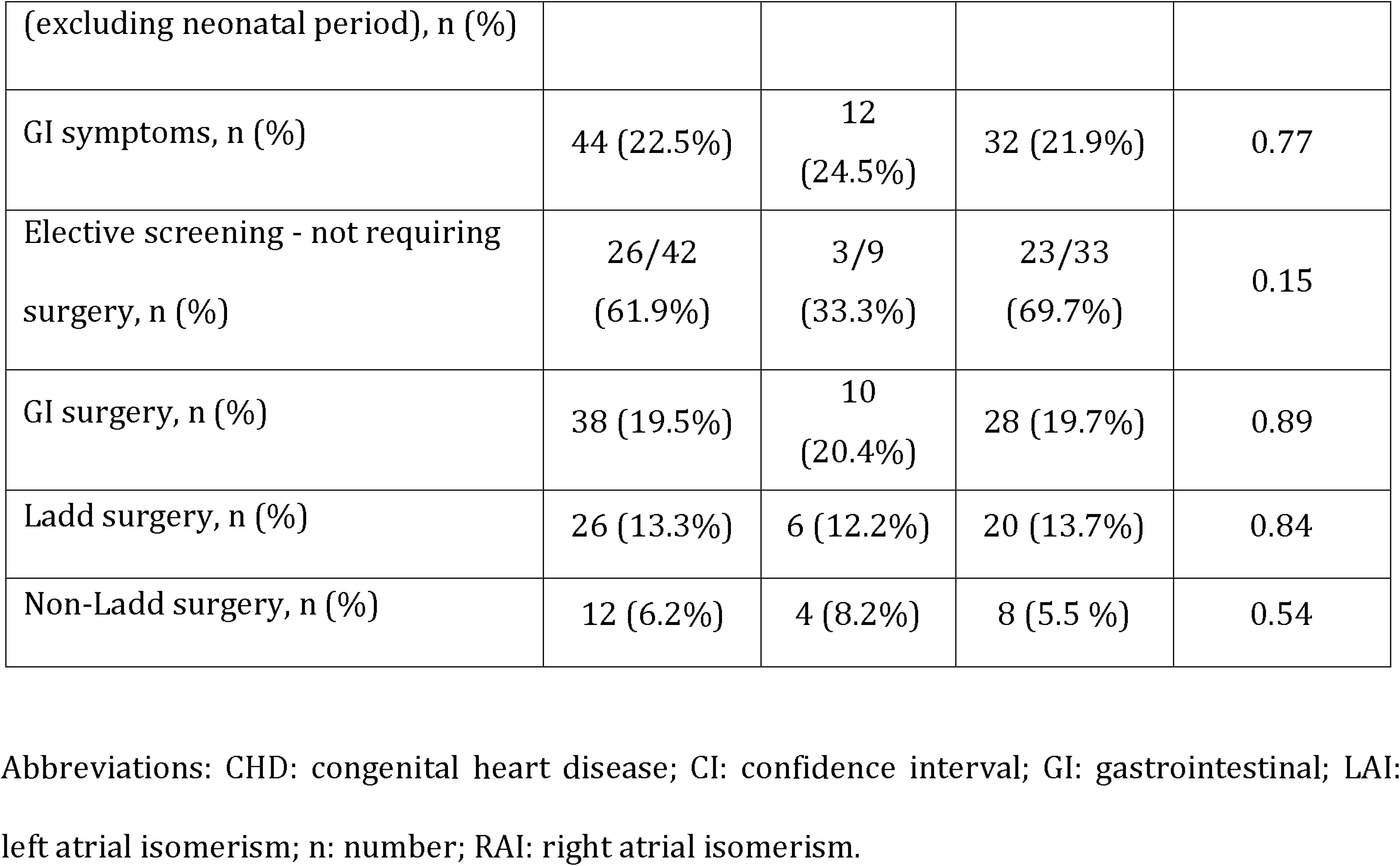
Demographic data and information related to heterotaxy, gastrointestinal symptoms, investigations and surgery.

### GI symptoms

Abdominal symptoms were recorded in nearly one-quarter of patients (44/195, 22.5%). In the majority (29/44, 65.9%), symptoms developed during the first year of life. Four cases of duodenal atresia were diagnosed prenatally and for all diagnosis was postnatally confirmed.

### GI investigations

Abdominal investigations to rule out rotational abnormalities were carried out in 42 of 195 (21.5%; 95% CI = 16.0-27.8%) patients. Twenty-two symptomatic patients were investigated. 12 had malrotation, but in two cases, surgery was not performed as patients died due to associated CHD. In eight patients, malrotation was excluded, however two did have surgery due to persistent symptoms. Two additional patients had surgery, but the results of the investigations were not recorded.

Of the 20 asymptomatic patients that were investigated, none was found to have malrotation.

### Surgery (GI)

From our total cohort, 38 patients (19.4%) had abdominal surgery (Table 1), 33 were symptomatic. Of the 38 cases: 17 (44.7%) underwent the Ladd procedure only; nine (23.7%) had both Ladd and non-Ladd abdominal surgeries and 12 (31.5%) had non-Ladd surgery (Table 2). In the 49 patients with RAI, 10 (20.4%) had abdominal surgery. This was also required in 28 of the 144 with LAI (19.4%) (Table 1 and 2).

A total of 27 non-Ladd surgical procedures were performed in 21 patients. Nine patients had duodenal or intestinal atresia, six required gastrostomy insertion or gastropexy, six had Nissen’s fundoplication and four procedures were performed to treat biliary atresia and gallbladder pathology.

**Table 2:**
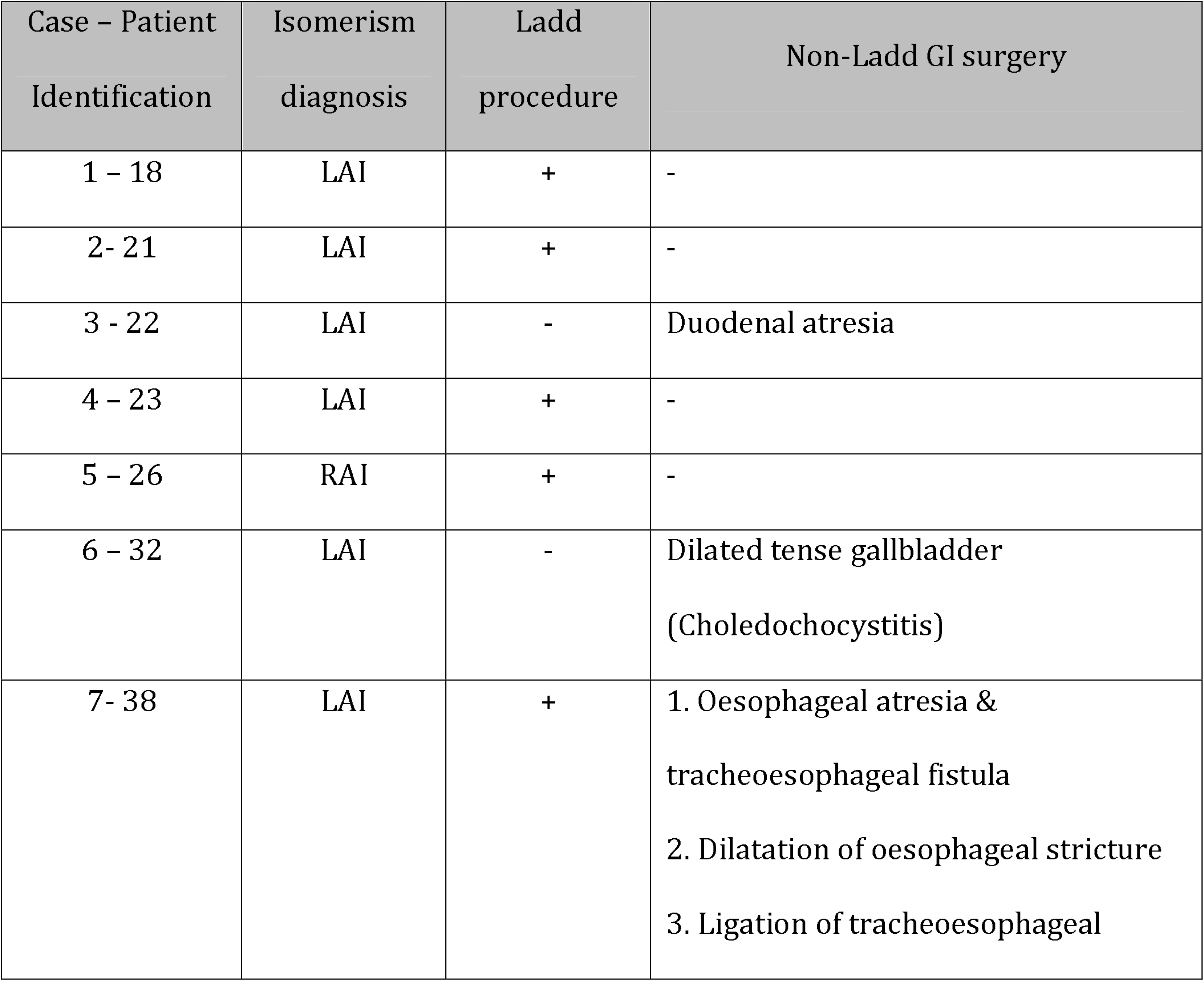
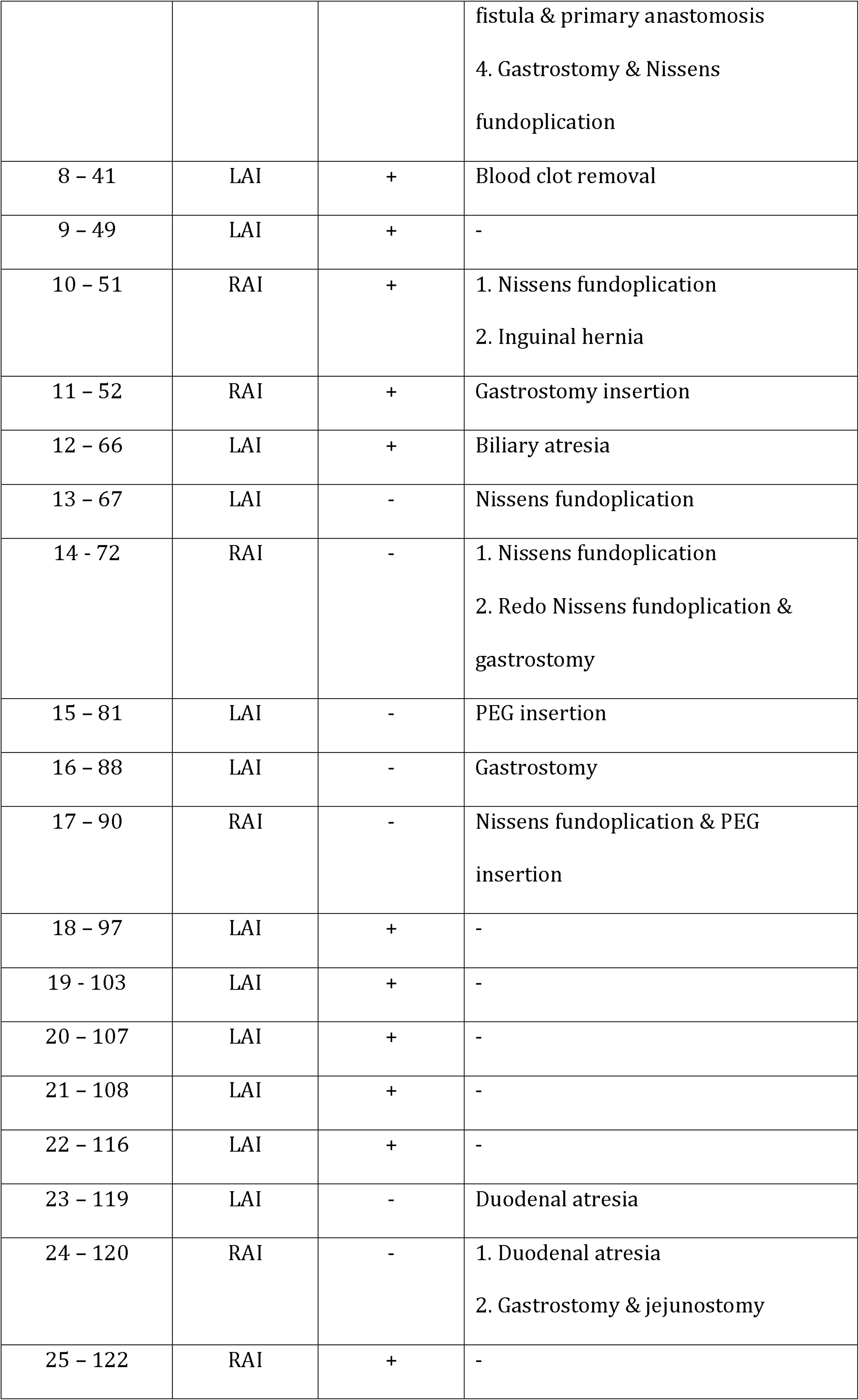
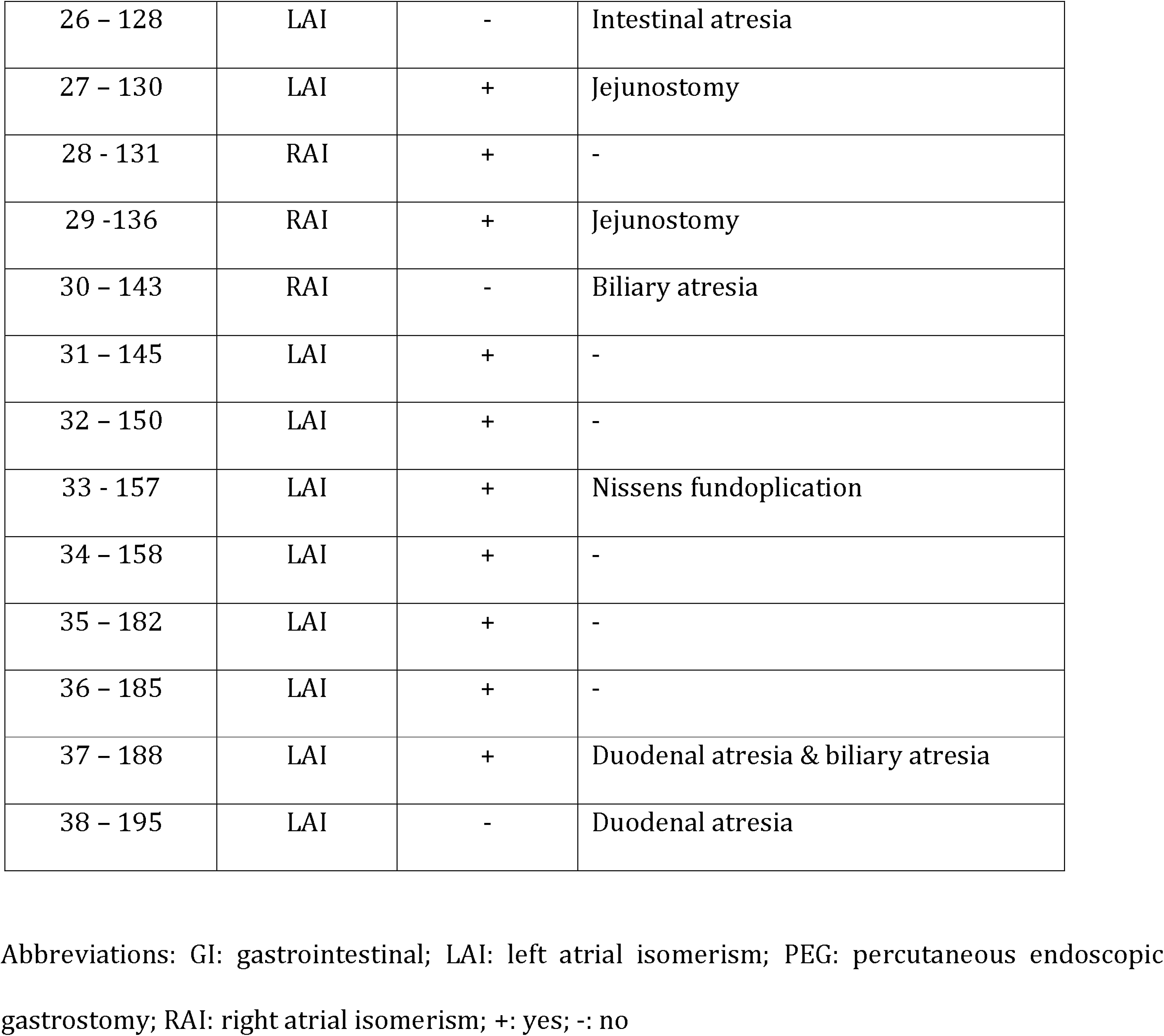
Data regarding detailed description of the abdominal surgeries performed in right and left isomerism patients.

### Follow up and outcome (KM survival curves)

Of the initial cohort, 161 patients were alive. The median follow-up time is 10.2 years (IQR =3.1 - 25.8). The median survival for patients without the need for cardiac intervention was 2.9 years, (IQR = 0.25 - 20.3). Figures 1a and 1b are Kaplan-Meier curves, showing freedom from cardiac intervention for the whole cohort and comparison between RAI and LAI, respectively. Ten-year freedom from cardiac surgery was 34% (95% CI = 27-42%) for the whole cohort, 7% (95% CI = 1-21%) for RAI and 42% for LAI (95% CI = 33- 51%), p < 0.0001.

Figures 2a and 2b represent Kaplan-Meier curves for freedom from any GI surgery for the whole cohort and comparison of RAI and LAI cohorts, respectively. Freedom from any GI surgery for the whole cohort at 1 year was 86% (95% CI = 80-91%), for RAI 86% (95% CI = 71-93%) and for LAI 86% (95% CI = 79-91%), p=0.98. Freedom at 10 years was 80% (95% CI = 75-85%) for all patients, 77% (95% CI = 59-87%) and 81% (95% CI = 73-86%) for RAI and LAI respectively, p = 0.65.

**Figure 2a:**
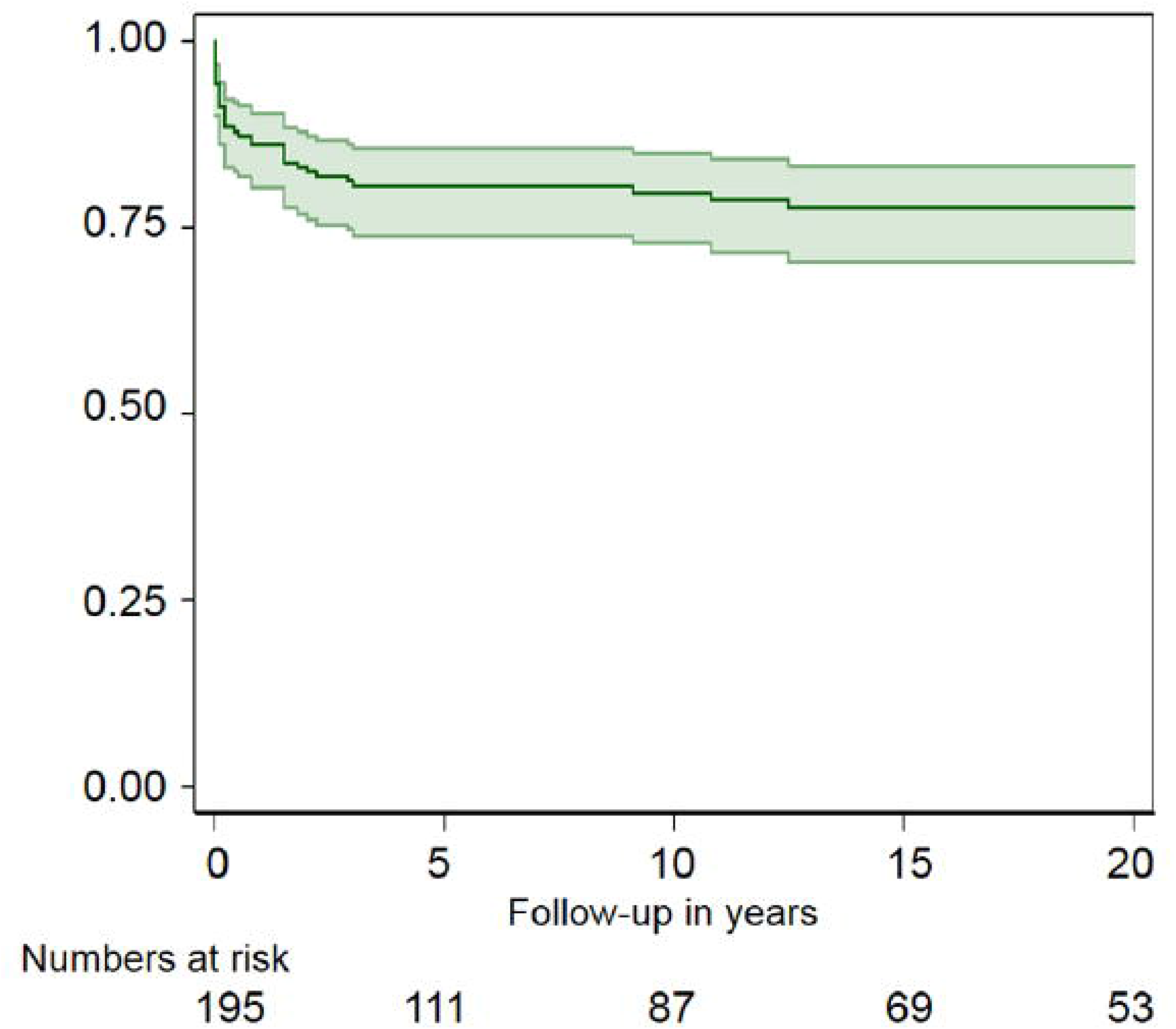
Kaplan-Meier curve, showing freedom from any GI surgery for the whole cohort.

**Figure 2b:**
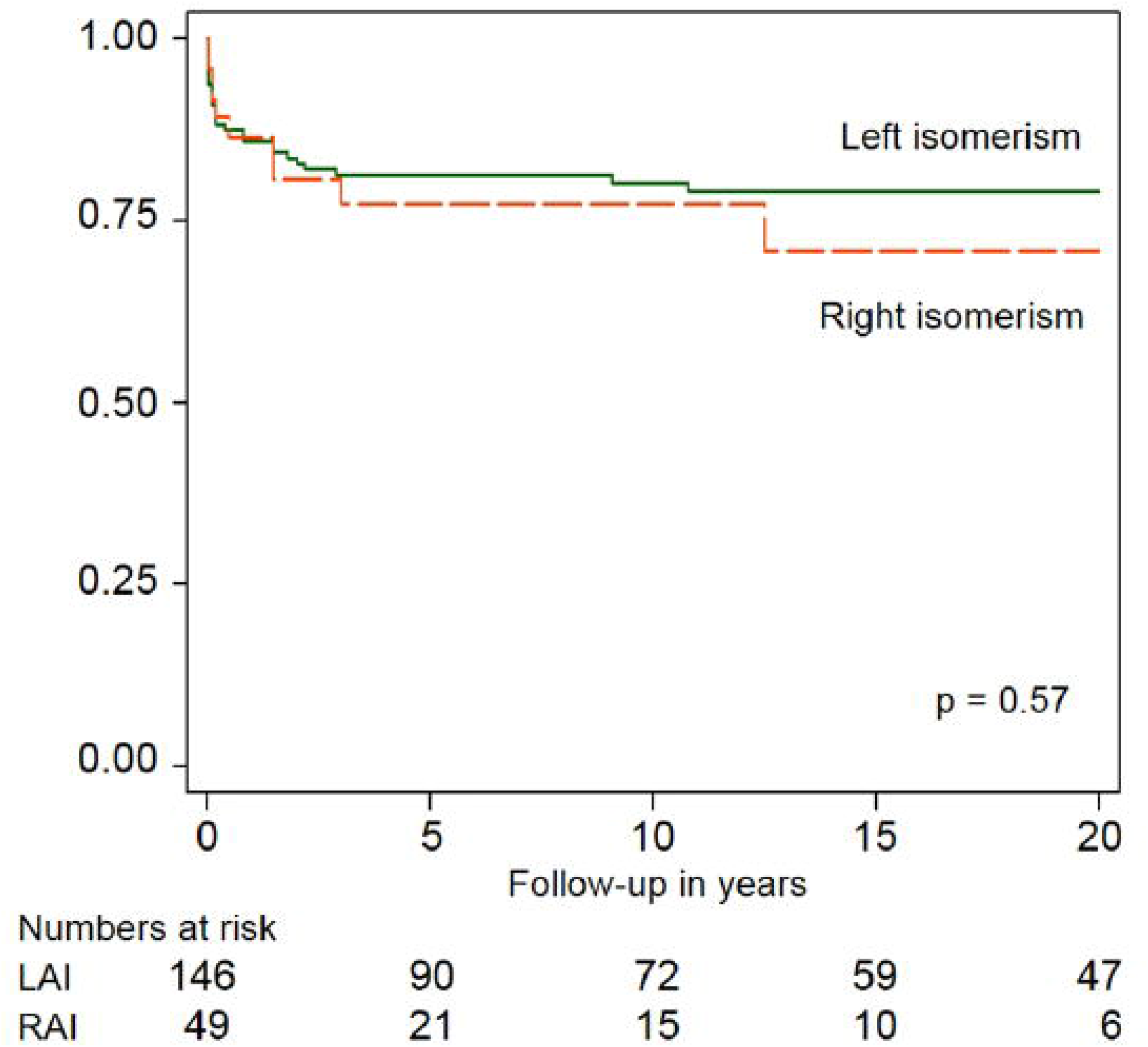
Kaplan-Meier curve, comparing freedom from any GI surgery for RAI versus LAI.

Figures 3a and 3b show freedom from Ladd procedure for the whole HS population and comparison of RAI and LAI cohorts, respectively. Overall freedom from Ladd procedure at 20 years was 86% (95% CI = 79-90%), for RAI 87% (95% CI = 72-94%) and LAI 85% (95% CI = 78-91%), p = 0.82.

**Figure 3a:**
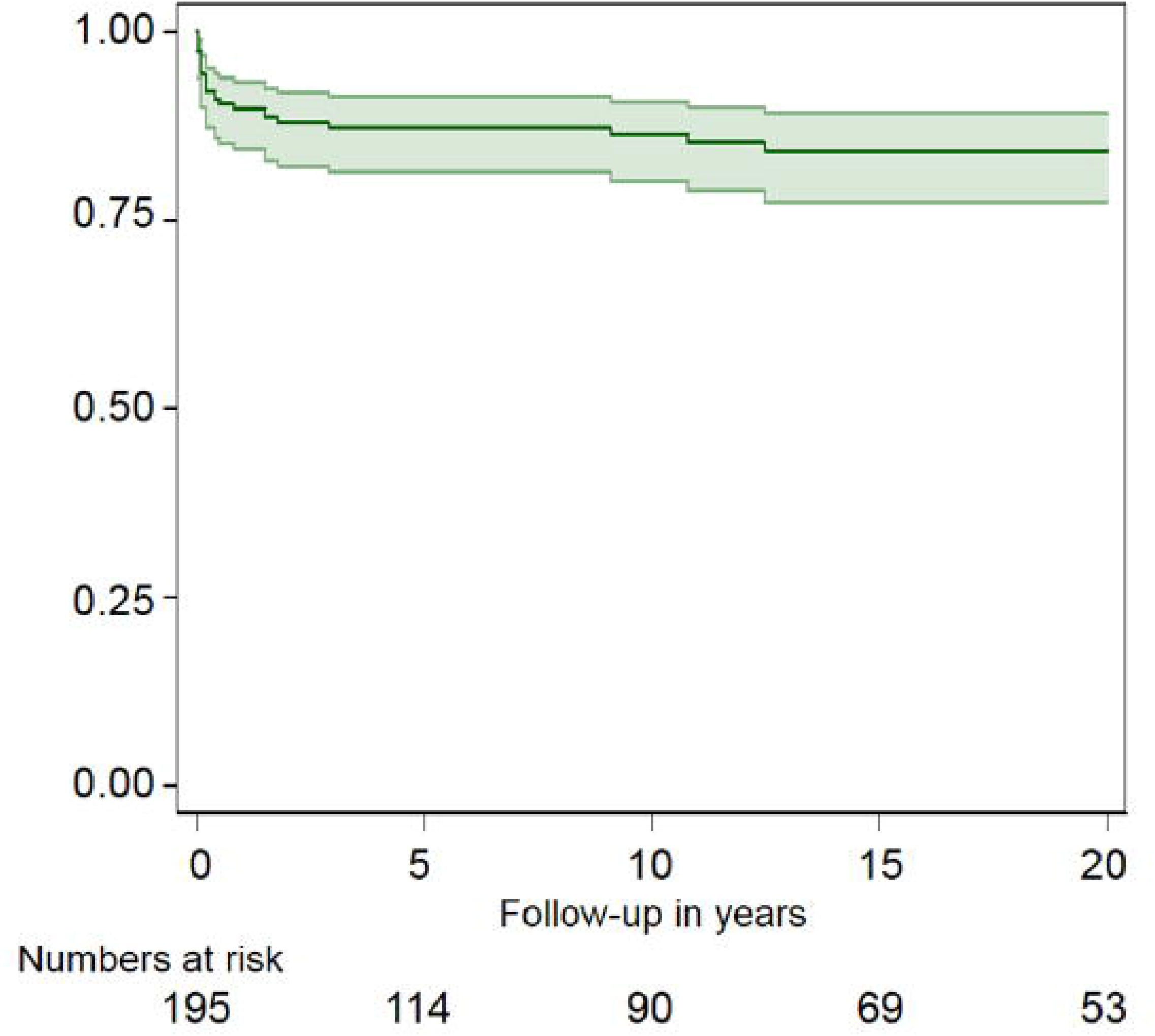
Kaplan-Meier curve, showing freedom from Ladd surgery for the whole cohort.

**Figure 3b:**
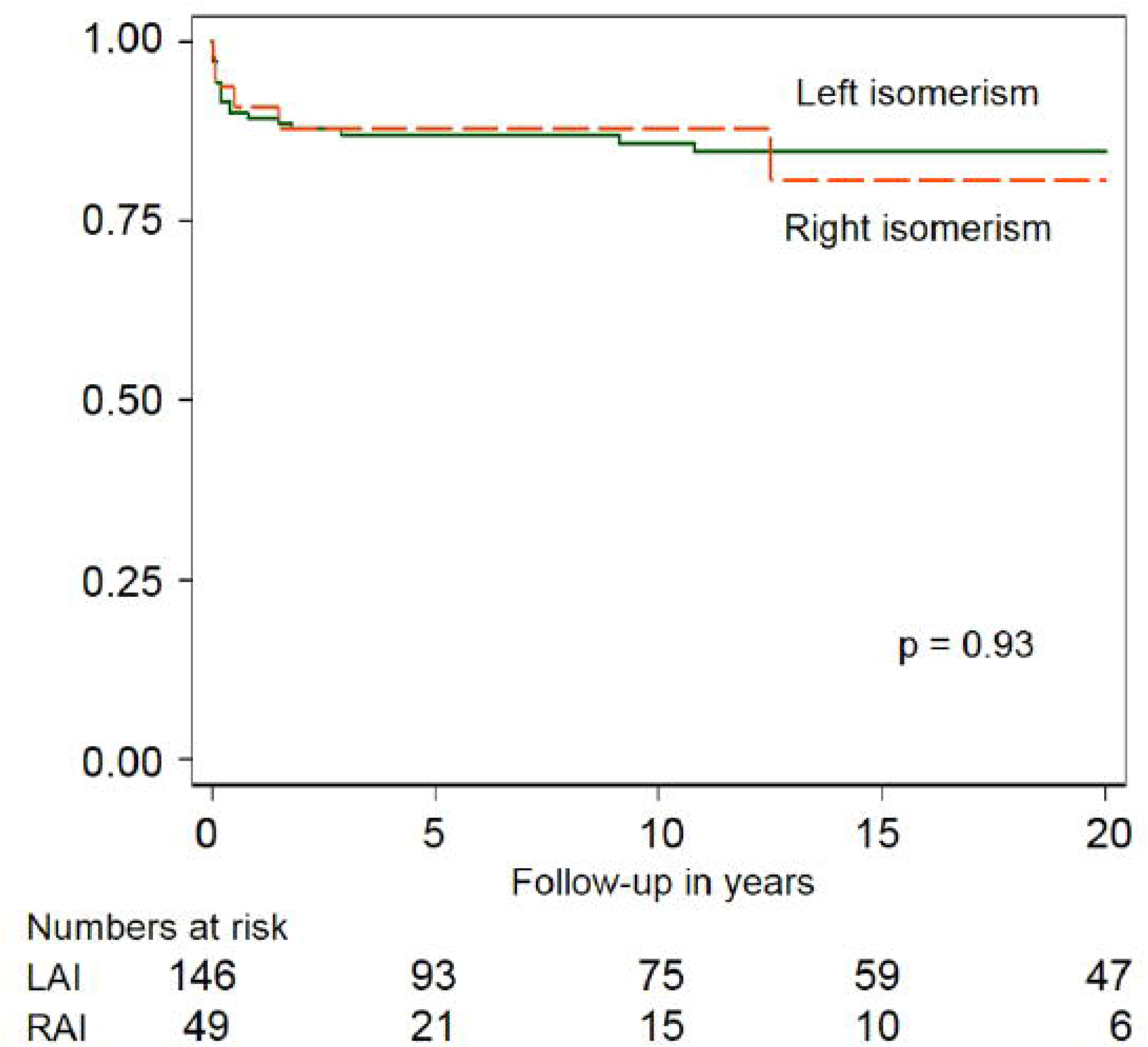
Kaplan-Meier curve, comparing freedom from Ladd surgery for RAI versus LAI.

## DISCUSSION

Abnormalities in intestinal rotation and fixation are well known to occur in conjunction with disordered cardiac looping found in heterotaxy. Establishing laterality during gastrulation at the beginning of the third week of development is essential for normal heart and gut development, because it specifies cells contributing to and patterning the right and left side of the heart and the rotation and fixation of the gut.^16, 17^ Typical malrotation refers to a spectrum of abnormal rotation of the bowel around the superior mesenteric vessels causing the small bowel to be prone to twist leading to midgut volvulus.

In our cohort of patients with RAI and LAI with a median follow up time of 10 years most patients required cardiac intervention and approximately 20% required GI surgery (Ladd procedure for classic malrotation in about two-thirds, non-Ladd interventions in about half of the cases, including intestinal atresia in 43%). There was no significant difference between the RAI and LAI cohorts with regards to GI comorbidity. Freedom from any GI surgery at 10 years was 80% and did not result in further GI comorbidities, reflecting low impact on cardiac outcome.

In our series, most patients had no GI related symptoms. Of the symptomatic group about two-thirds developed symptoms within the first year of life. In the literature, up to 40% of patients with malrotation present within the first week of life and up to 75% to 85% have been diagnosed by age 1 year. The diagnosis of the remaining 15% to 25% of patients is usually made during childhood but can be into late adulthood. However, it is difficult to estimate the true incidence of malrotation in the adult population.^4, 8, 14, 15^In our series, patients requiring Ladd procedure were mostly symptomatic (25/26).

The incidence of non-Ladd abdominal surgery in our cohort is probably also an indication that disordered looping in intestines comprises a wider spectrum of functional abnormalities than typical malrotation.

In our series, there was no patient presenting with acute mid-gut volvulus. This is in agreement with a review by Elder and colleagues.^18^ In their series, which included 99 HS patients, 23 had Ladd’s procedure, only one presented with acute abdomen, but no volvulus was found at surgery. Of the remaining 76 patients who did not undergo a Ladd procedure, 14 had normal intestinal anatomy, 5 had malrotation, and 57 were never evaluated for intestinal malrotation. Average follow-up time was 5.1 years. It is concluded that HS patients who did not have screening and prophylactic surgery were not exposed to a significant risk of midgut volvulus ^18^.

The management GI strategy for the asymptomatic HS patient has been the subject of academic debate in the literature over the past decade.^5, 19, 20^ Advances in cardiac care result in HS patients reaching cardiac stability^19^ with asymptomatic intestinal rotation.^21^

There is no agreement concerning screening for malrotation in HS patients.^5^ Papillon and colleagues doubt the true clinical significance of the findings of malrotation in the upper GI tract contrast study in an asymptomatic HS patient.^22^ In a recent series, they noted that of 30 asymptomatic patients with investigation findings of duodeno-jejunal malposition - atypical malrotation, none of them had true malrotation or a narrow mesentery at the time of abdominal exploration.^22, 23^ They also observed 138 asymptomatic patients without imaging and none developed volvulus. ^11, 22^ White et al found that the presence of malrotation does not impact the outcomes following cardiac surgery for patients with complex CHD. The incidence of volvulus in this group is also low and therefore does not advocate for a prophylactic Ladd procedure.^24^

The concerns regarding Ladd procedure itself are related to the predisposition for post-operative complications, particularly small bowel obstruction with rates ranging from 3-15%^25^ and controversy exists regarding possible higher morbidity when performed in asymptomatic HS patients versus (vs) the symptomatic ones (22% vs 13%).^12^ However, the risk of not operating on an asymptomatic abnormality of intestinal rotation is the morbidity associated with potential midgut volvulus and an emergency abdominal surgery.

Yu and colleagues in a retrospective review support prophylactic Ladd procedure in all asymptomatic HS patients with suspected malrotation,^5^ as they encounter similar rates of complications and hospital stay after Ladd procedure between HS and non-HS patients with presumed malrotation. A similar argument supporting prophylactic Ladd procedure has been published by other groups^10, 11, 25^

An alternate opinion has been presented by Choi and colleagues who studied 177 HS patients, 152 were asymptomatic and nine had screening, six with abnormal results.^26^ A total of 143 were followed-up, only four developed symptoms and only one developed malrotation requiring Ladd procedure. The incidence of volvulus was 0.6%, lower than the rates of small bowel obstruction published elsewhere and none needed bowel resection. Another retrospective study of over 300 patients with HS and intestinal rotation abnormalities from 41 tertiary children’s hospitals found that Ladd procedure was associated with a 120% increase in the odds of major morbidity and mortality, even after considering various risk factors.^4^ Therefore, close observation, might reduce the odds of future complications in this patients group.^4^ Equivalent postoperative outcome has been shown between prophylactic and emergent Ladd procedures, and complications occurred more frequently in patients who had a Ladd procedure rather than in patients who were observed, suggesting non-operative management as a reasonable alternative to prophylactic surgery.^28^ In a recent systematic review another group concludes that given the spectrum of intestinal rotation abnormalities, ranging from absence of normal rotation, which is relatively benign, to severe malrotation that carries higher risk of obstruction and midgut volvulus and associated cardiac comorbidity, asymptomatic patients should be deferred from an unnecessary Ladd procedure.^29^ This view is supported by others.^4, 14, 26, 27, 30^

In our study group, there were no major GI surgical complications, but no patient was treated prophylactically. None of our patients required emergency Ladd procedure because they did not present with acute midgut volvulus.

Limitations of our study include the retrospective review of the pediatric cardiology notes, particularly due to occasional brief reports on abdominal symptomatology and complications after GI procedures. We tried to overcome this limitation, by including relevant questions on the symptomatology and on the surgical complications in the questionnaires we sent to parents. For patients who died from CHD in the neonatal period, autopsy findings to confirm or not the presence of malrotation are not available. A larger, prospective series is needed to determine if these theories hold true.

## Conclusions

In our cohort, one in five patients required abdominal surgery, the majority in their first year, similar in RAI and LAI. Between one and ten years of follow up the impact of GI abnormalities on outcome was minimal. Medium term survival was related to CHD.

## Data Availability

Extra data is available by emailing AF. Anastasia Fotaki

**Abbreviations**
CHD: congenital heart disease;
CI: confidence interval;
GI: gastrointestinal;
HS: heterotaxy syndrome;
IQR: inter-quartile range;
LAI: left atrial isomerism;
n: number;
PEG: percutaneous endoscopic gastrostomy;
RAI: right atrial isomerism;
SVC: superior vena cava

## ACKNOWLEDGEMENTS

Debbie Rolfe, Senior Regulatory Assurance Manager, St. George’s University Hospitals NHS Foundation Trust, for guidance with Integrated Research Application System and Research Governance.

Medical Records Department, Royal Brompton & Harefield NHS Foundation Trust, for assistance in locating and retrieving paper notes.

## DATA SHARING STATEMENT

Extra data is available by emailing AF.

## CONTRIBUTORY STATEMENT

AF and VD were involved in the retrieval of patient records and data collection. AF, JSC, SB and VD were involved in the design of the questionnaires and letters to parents. AF, WB, JSC were involved in the statistical analysis of the data. All authors were involved in the conception, design and interpretation of the data. All authors have been involved in drafting the manuscript or revising in critically and have given final approval of the version to be published. AF is responsible for the overall content as guarantor together with JSC.

## AFFILIATIONS

Brompton Centre for Fetal Cardiology, Royal Brompton & Harefield NHS Foundation Trust, Royal Brompton Hospital, London, UK (A.F., V.L.D., J.S.C.); Fetal Medicine Unit, St. George’s University Hospitals NHS Foundation Trust, St. George’s Hospital, London, UK (A.F., J.S.C.); Department of Medical Statistics, Research & Development, Royal Brompton & Harefield NHS Foundation Trust, London, UK (W.B.); Department of Neonatal & Paediatric Surgery, St. George’s University Hospitals NHS Foundation Trust, London, UK (S.G.); Department of Neonatal Unit, St George’s University Hospitals NHS Foundation Trust, London, UK (S.B.); Molecular & Clinical Sciences Research Institute, St George’s University of London, London, UK (J.S.C.).

